# Comprehensive Analysis of Mitochondrial DNA Variation in the Taiwan Biobank: Implications for Complex Traits and Population Genetics

**DOI:** 10.1101/2024.10.28.24316086

**Authors:** Ting-Hsuan Chou, Pei-Miao Chien, Pin-Xuan Chen, Ni-Chung Lee, Hurng-Yi Wang, Yi-Cheng Chang, Pei-Lung Chen, Jacob Shu-Jui Hsu

**Affiliations:** Graduate Institute of Medical Genomics and Proteomics, National Taiwan University College of Medicine, Taipei, Taiwan; Department of Medical Genetics, National Taiwan University Hospital, Taipei, Taiwan; Department of Pediatrics, National Taiwan University Hospital, Taipei, Taiwan; Department of Pediatrics, National Taiwan University College of Medicine, Taipei, Taiwan; Graduate Institute of Clinical Medicine, College of Medicine, National Taiwan University, Taipei, Taiwan; Genome and Systems Biology Degree Program, National Taiwan University and Academia Sinica, Taipei, Taiwan; Institute of Ecology and Evolutionary Biology, National Taiwan University, Taipei, Taiwan; Institute of Biomedical Sciences, Academia Sinica, Taipei, Taiwan; Department of Internal Medicine, National Taiwan University Hospital, Taipei, Taiwan

## Abstract

This study presents a comprehensive analysis of mitochondrial DNA (mtDNA) variation in the Taiwan Biobank (TWB), providing insights into the genetic characteristics of the Taiwanese population and the implications of mtDNA in complex traits. We performed mtDNA genotyping on 1,492 individuals using whole-genome sequencing data and imputed mtDNA variants for 101,473 participants from microarray data. Our analysis identified 23 confirmed pathogenic mtDNA variants, with approximately 1 in 180 individuals carrying such variants. Further exploration of mtDNA haplogroups and ancestry revealed no direct correlation between nuclear and mitochondrial genomes, which reflects their distinct inheritance patterns and evolutionary histories. In a mitochondrial genome-wide association study across 86 traits and 306 mtDNA variants, we discovered novel associations between *MT-ND2* gene variants and high myopia, as well as 14 mtDNA variants linked to renal function biomarkers. Notably, renal-associated variants clustered into two main groups: ancestral variants of macrohaplogroup M associated with poorer renal function and variants of the B4b sub-haplogroup linked to improved renal function markers. Our findings highlight the importance of population-specific genetic studies, contributing to our understanding of mitochondrial genetics in the Taiwanese population and its implications for health and disease.

## Introduction

Mitochondria, often referred to as the “powerhouse” of cells, are double-membrane organelles presented in almost every eukaryotic cell and serve as the primary source of cellular energy production through ATP synthesis via oxidative phosphorylation (OXPHOS)^1^. Each mitochondrion contains its own genome (mtDNA), a circular double-stranded DNA molecule. In humans, the mitochondrial genome spans 16,569 base pairs and contains 37 genes encoding 13 protein subunits of the OXPHOS system, 22 tRNAs, and two rRNAs, all of which are crucial for its maintenance and expression^2,3^.

Mitochondrial DNA exists in multiple copies within each cell, with the number of copies varying among individuals, tissues, and cells based on metabolic demands^4^. Due to multiple copies of mtDNA, accumulated variants can manifest in two forms: homoplasmy and heteroplasmy. Homoplasmy occurs when a specific variant is uniformly present across all mtDNA copies within an individual, while heteroplasmy arises when a mixture of different molecules exists. Since mtDNA is maternally inherited^5^ and lacks intermolecular recombination^6^, variants within a population are grouped into mtDNA haplotypes, commonly called haplogroups. These haplogroups have proven invaluable for tracking human biogeography^7^.

Given the central role of mitochondria in energy production, it is not surprising that mtDNA variants have been associated with various diseases, including type 2 diabetes ^8^, cardiomyopathy^9^, renal disease^10^, and Parkinson’s disease^11^. Recent population-level studies have further elucidated the influence of mtDNA variants on human health. For instance, research conducted within Biobank Japan involving 1,928 individuals revealed pleiotropy of mtDNA genetic risk on the five late-onset human complex traits such as creatine kinase^12^. Similarly, a larger investigation using the UK Biobank with 358,916 participants identified new associations between mtDNA variants and various traits, including type 2 diabetes and markers of liver and kidney function^13^. These studies underscore the critical role of common mtDNA variations in shaping complex human traits.

The Taiwan Biobank (TWB), established in 2012, is a pivotal genetic research resource. This prospective study has recruited over 200,000 individuals, generating a comprehensive repository of genomic and phenotypic data. The extensive data encompasses participants’ demographics, socioeconomic status, family history, and self-reported disease profiles. Given that the majority of Taiwanese participants are of Han Chinese ancestry, analyses from TWB can offer unique insights into the mtDNA impacts on health in Taiwanese and other East Asian populations^14,15^.

In this study, we comprehensively dissect the mitochondrial genetic profiles of the Taiwanese population utilizing data from the TWB. Our investigation had three primary objectives. First, to characterize the spectrum of mtDNA variation in the TWB, we implemented the GATK mitochondrial variant calling pipeline to accurately identify both homoplasmies and heteroplasmies from 1,492 WGS samples. Second, to explore the mitochondrial genetic structure and ancestry patterns, we performed Principal Component Analysis (PCA) on mtDNA variants and assessed the correlation between haplogroup classifications and their ancestral origins. Finally, to investigate the role of mitochondrial variation in complex traits, we accurately imputed mtDNA variants for 101,473 individuals using microarray data by building a Taiwanese-specific mitochondrial reference panel based on WGS data. This imputation enabled us to conduct a hypothesis-free mitochondrial-wide association study involving 306 mtDNA variants and 81 complex traits. Through the analysis, we aim to uncover significant associations between mtDNA variations and various health outcomes, thereby contributing valuable insights into the role of mitochondrial genetics in complex traits within the Taiwanese population.

## Methods

### Data source

The research cohort was drawn from the Taiwan Biobank (TWB), a prospective study that has recruited over 200,000 individuals. The TWB provides extensive phenotype data, including demographics, socioeconomic status, environmental exposures, lifestyle factors, dietary habits, family history, and self-reported diseases gathered through structured questionnaires. Additionally, anthropometric measurements, including blood and urine samples, were obtained at the time of enrollment for subsequent biomarker analysis. For this study, we utilized a total of 1,492 whole-genome sequencing (WGS) data and 120,163 microarray data sourced from the TWB with ethical approval (TWBR11106-05 and 202108074RINC).

### Mitochondrial DNA variant calling from the WGS data

In the TWB dataset, 1,492 WGS samples were sequenced by using HiSeq 2500 (n=555), HiSeq 4000 (n=634), or NovaSeq 6000 (n=303), which achieved high depth on autosomal chromosomes (35-45x). To identify mtDNA variants, we employed the GATK Best Practices for SNP/Indel Variant Calling in Mitochondria (V.4.1.8.0)^16^. In brief, the reads were aligned to GRCh38 using BWA-MEM (version 0.7.17), which includes the revised Cambridge Reference Sequence (rCRS, NC_012920.1) serving as the mitochondrial reference genome. A double alignment strategy was conducted to align the control region and the other region separately; this approach was necessary due to the circular nature of the mitochondria’s genome. The caller used Mutect2 in mitochondria mode to detect variants with low variant allele frequency (VAF).

Subsequent stringent filtering steps were applied following GATK best practices for mitochondrial variants. At the sample level, we filtered samples with low mitochondrial copy numbers (<50) and excluded samples with contamination greater than 0.02. For genotype calls, we filtered out variants with a VAF of less than 0.1. At the variant level, we removed alleles found in regions where the sequence context makes it difficult to distinguish true variants from technical artifacts, including artifact-prone regions in six mtDNA positions (301, 302, 310, 316, 3107, 16182) and indels were only present as multi-allelic calls across all samples, as well as alleles for which no sample had a pass genotype.

### Haplogroup assignment

We utilized high-quality mtDNA variants from WGS as input and classified each individual into a mitochondrial haplogroup by HaploGrep (v2.4.0)^17^ based on the revised tree Phylotree17_FU1^18^. The first letter of the haplogroups was defined as macrohaplogroup. Each haplogroup is phylogenetically associated with three origins: African, Asian, and European, as described in MITOMAP^19^.

### Principal Component Analysis (PCA) on the nuclear DNA (nDNA) and mtDNA variant

To explore clustering patterns of autosomal and mitochondrial genetic structure, we employed Principal Component Analysis (PCA) as a linear dimensionality reduction technique. The autosomal variant detection was conducted as previously described^20^. Individuals with missing rates >0.02 were excluded from the analysis. Nuclear PCAs were analyzed, focusing on a subset of SNVs adhering to the following criteria: SNVs with a VQSR tranche <99.7 and genotype call rate >0.98 biallelic variants. We then conducted PCA analysis using PLINK to elucidate the underlying clustering pattern^21^.

Information regarding individuals’ places of origin was obtained from a self-reported questionnaire, which included data on both maternal and paternal ancestries. Based on this data, individuals from the TWB were classified into four main clusters: “Holo,” “Hakka,” “Southern Han Chinese,” and “Northern Han Chinese.” The categorization of “Southern Han Chinese” and “Northern Han Chinese” was based on their respective provincial regions in relation to the Yangtze River.

### Quality control of mtDNA variants from the microarray data

We utilized 120,163 microarray data from the TWBv2 SNP array, where participants were genotyped using the Thermo Fisher Scientific Axiom Genome-Wide TWB 2.0 Array. This genotyping array includes 752,921 probes designed to assay 686,463 SNVs, as well as 815 mtDNA variants. The analysis was performed using officially released PLINK files, which were processed with the Axiom Analysis Suite, following the manufacturer’s recommended best practice workflow.

To ensure high-quality mtDNA variant data, we applied stringent quality control (QC) criteria. We excluded individuals with a sample missing rate >0.02 and variants with a missing rate >0.02. Additionally, we compared allele frequencies (AFs) observed in the microarray with those from WGS. Variants with AFs deviating more than 0.2 from WGS observed frequencies were removed from the subsequent analysis.

To evaluate the detection limit of heteroplasmies under standard procedures, we leveraged data from 1,426 individuals who had both WGS and microarray data available. We compared the genotype calls between the array and WGS datasets for variants that had designed probes and were detected in the WGS datasets to assess consistency. Specifically, we used the WGS call set as the truth set, allowing us to compare the detection rates of the array data across different VAF bins with those detected by WGS. This analysis provides a thorough evaluation of the microarray’s capability to detect heteroplasmies under different VAFs. As depicted in Figure S4B, since the detection of heteroplasmies is rare in the genotyping of mtDNA variants, we set all the heteroplasmies as missing in WGS when constructing the mitochondrial imputation reference panel.

### Evaluation of imputation accuracy

Before initiating imputation, we assessed the feasibility and accuracy of imputing mtDNA variants using a sample of 1,426 individuals who had both WGS and microarray data available. These individuals were divided into two groups: 1,000 formed the reference set, and the remaining 426 constituted the test set. We then constructed an mtDNA imputation reference panel from the WGS sequences of the 1,000 individuals and imputed mtDNA variants from the microarray data of the 426 test individuals.

The mtDNA scaffold haplotypes were pre-phased using SHAPEIT2^22^ before imputation to ensure compatibility with the IMPUTE2 algorithm. Imputation of mtDNA variants was conducted by IMPUTE2^23^, covering the full mitochondrial genome with region boundaries set to –int 1 16579. Given the absence of an available mitochondrial genetic map, we created a genetic map indicating little to no recombination of the mitochondrial genome.

We evaluated the imputation performance in both haploid and diploid settings and observed no significant differences. After imputation, we assessed genotype concordance between the WGS and imputed variants for each AF bin separately (0-0.005, 0.005-0.01, 0.01-0.05, 0.05-0.1, 0.1-0.2, 0.2-0.3, 0.3-0.4, 0.4-0.5, 0.5-0.6, 0.6-0.7, 0.7-0.8, 0.9-1.0). This analysis enabled us to establish post-imputation filtering criteria, setting a threshold INFO score of 0.7 to ensure high confidence in the imputed data.

### Imputation of microarray data

We constructed a Taiwanese-specific mitochondrial reference panel from 1,465 WGS sequences. After applying pre-imputation filtering, our dataset included 101,473 unrelated EAS samples and 563 mtDNA variants that were biallelic, had a missing rate of less than 0.02, and exhibited AF deviations from WGS of less than 0.2. Post-imputation QC filtering was applied to variants with MAF >0.01 and an imputation INFO score >0.7. This process resulted in 306 high-quality mtDNA variants, which demonstrated high allele frequency consistency with WGS data (Pearson r = 0.999), suitable for subsequent association analysis.

### Preparation of phenotypic traits for analyses

We gathered phenotypic data on disease status via questionnaires and clinical measurements from the TWB. We included self-reported diseases with at least 100 cases, resulting in a selection of 48 diseases. For quantitative traits, these measurements encompass seven categories, offering a comprehensive assessment of participant health. These categories include anthropometric measurements (n = 7), lung function (N=3), bone density (n=2), cardiovascular function (n=3), hematological parameters (n=5), metabolism (n=6), liver (n=6) and kidney function (n=6). We removed individuals whose measurements deviated by more than five standard deviations from the mean to eliminate outliers and applied rank inverse normal transformation (RINT) to standardize each phenotype.

### Mitochondrial genome-wide association analysis

Initially, we defined a subset of unrelated East Asian (EAS) individuals to be included in the downstream association analysis. We inferred a genetically EAS group from the TWB using 1000 Genomes (1KG) phase 3 samples as the population reference panel^24^. PCA was performed on common autosomal variants between TWB and 1KG datasets. The variants selected were biallelic with a missing rate <0.02, *r*^2^ <0.1, MAF >0.05, and located outside long-range LD regions (chr6: 25-35Mb, chr8: 7-13Mb). Then, we used the Random Forest classifier to predict the EAS group^15^.

To ensure the analysis was conducted on unrelated individuals, we calculated kinship coefficients using KING^25^ based on autosomal variants. Individuals identified with a kinship coefficient greater than 0.0884, indicative of second-degree relatives or closer relationships, were excluded from the study.

For the statistical analysis, we implemented an additive model and conducted regression analyses using the glm() function in R, tailored for quantitative traits (linear model) and binary traits (logistic model). These analyses were adjusted for age, sex, age and sex interaction, the top 10 nuclear PCs, and genotyping batches to mitigate potential confounding factors. For sex-specific phenotypes, adjustments were made for age and the top 10 nuclear PCs. We applied a Bonferroni correction to set the significance threshold at P = 0.05/306 = 1.4 × 10^−4^. Additionally, considering the number of traits analyzed, we adopted more conservative thresholds of P = 0.05/306/48 *t*r*aits* = 3.4 × 10^−6^ for binary traits, and P = 0.05/306/38 *t*r*aits* = 4.3 × 10^−6^ for quantitative traits.

### Association of mtDNA haplogroups

Using imputed mtDNA genotypes, we applied Haplogrep2 to assign haplogroup for each individual ^17^. We applied filters before performing haplogroup assignments, including individual missing rate <0.02, genotype missing rate <0.02, and MAF >0.01. Following the haplogroup assignment, we analyzed the distribution of haplogroups and conducted PCA analysis on these mtDNA variants. The results from the genotyping array were compared with those in WGS to assess the similarity and accuracy.

Subsequently, we focused on the association between haplogroups and renal markers, including serum creatinine (Scr) and estimated glomerular filtration rate (eGFR). Both values were rank-inverse normal transformed. The association analysis was restricted to haplogroups represented by at least 500 individuals to ensure sufficient statistical power. We employed GLM with a Gaussian distribution, adjusting for genotyping batches, age, and sex, and the most prevalent haplogroup M7b was set as the reference. We set the significance threshold at P < 1.22 × 10^−3^ (0.05/41 ℎ*aplog*r*oups*), calculated based on the number of haplogroups tested.

## Results

### mtDNA genotyping across 1,492 TWB individuals

In this study, we conducted mtDNA genotyping on 1,492 individuals from the TWB using the GATK pipeline for variant calling. This approach allowed us to characterize the spectrum of mtDNA variation in the TWB by identifying both homoplasmic and heteroplasmic derived from WGS data. Following rigorous filtering criteria, we retained 1,465 samples for further analysis. These samples exhibited nDNA coverage ranging from 35x to 45x, and mtDNA coverage ranged from 2500x to 3000x. The average mtDNA copy number across these samples was estimated to be 155^26^. (Figure 1A-1C).

**Figure 1.**
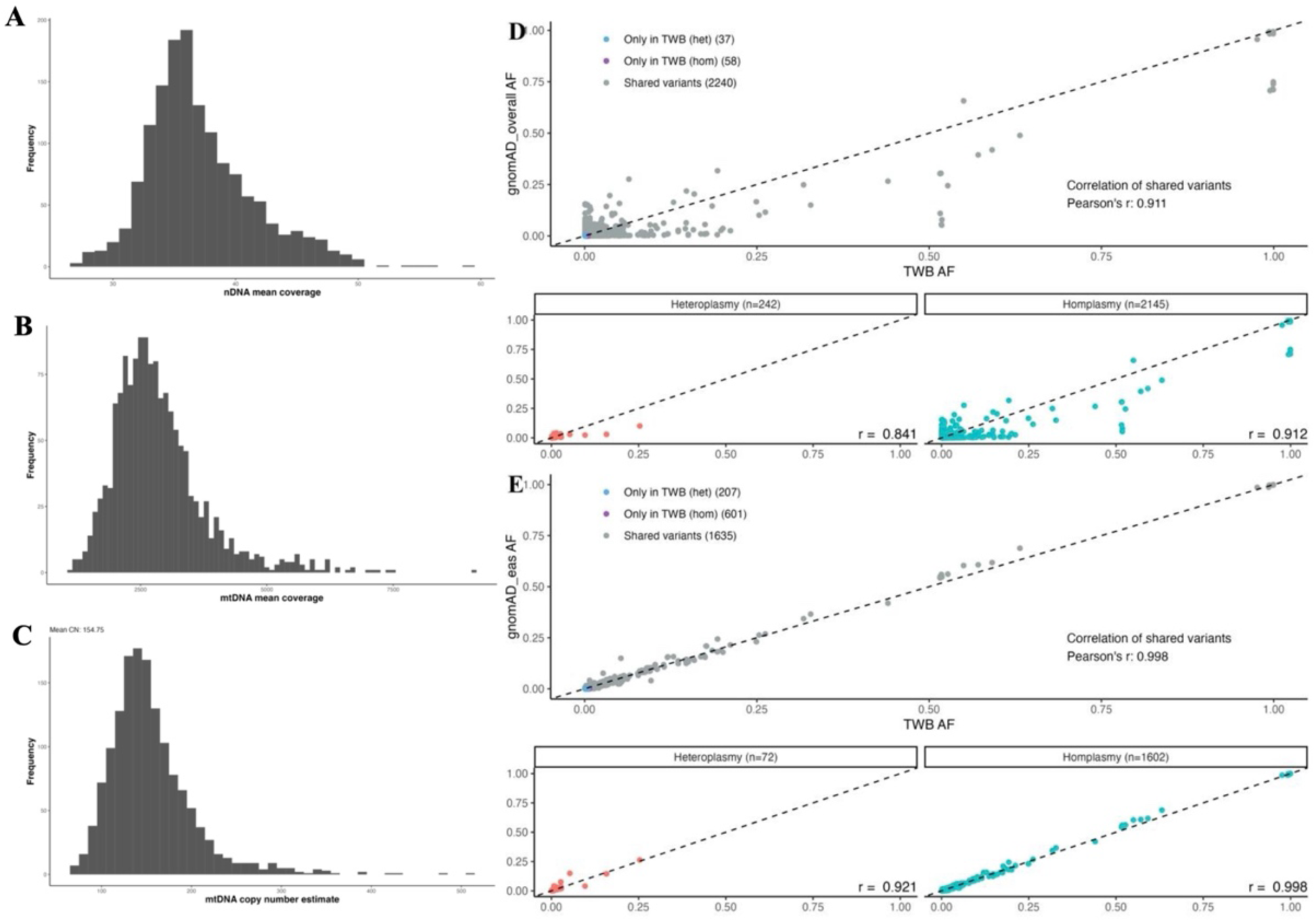
mtDNA variant statistics in Taiwan Biobank. (A-C) The histograms display nDNA coverage (A), mtDNA coverage (B), and mitochondrial copy number (mtCN) (C) across 1,465 WGS samples. (D) Scatter plots displaying correlations of allele frequencies between TWB and gnomAD: overall AFs (D), East Asian (EAS) AFs (E). Grey dots represent shared variants between the two datasets. Blue and purple dots denote heteroplasmies and homoplasmies exclusive to TWB. The line of identity is depicted in each plot, emphasizing the correlations. The subplots below compare heteroplasmic and homoplasmic variants separately.

Our analysis unveiled a total of 2,361 mitochondrial variants, consisting of 2,289 SNVs and 72 indels. The majority of SNVs were transitions rather than transversions, and most detected variants were found to be homoplasmic. Furthermore, our investigation of AF spectra revealed that over half of the identified variants occurred uniquely in one or two individuals, with the majority having AFs below 1% (Figure S1).

When comparing our TWB mtDNA variant call set with the gnomAD database (v3.1), the Taiwanese cohort exhibited a close genetic resemblance to gnomAD’s East Asian population, with a Pearson correlation of 0.998 for AF distributions of shared variants (Figure 1D and IE). However, about 30% of the variants found in TWB were not reported in the East Asian population of gnomAD, suggesting notable differences and underscoring the importance of population-specific studies.

### Capability of mtDNA and nuclear genetic structure in reflecting ancestral information

Due to the nature of maternal inheritance and lack of recombination, mtDNA haplogroups provide a framework for understanding the maternal lineage^7^. The Phylotree database offers a thorough phylogenetic tree of worldwide human mtDNA variation, comprising over 5,400 haplogroups with their defining mutations^27^. To discern the haplogroup within each individual, we used HaploGrep2 for haplogroup assignment^17^. Our analysis revealed that the most prevalent mtDNA haplogroups among the TWB participants were M, D, F, and B, all indicative of Asian ancestry and reflective of the genetic background of the Taiwanese population (Figure 2A and 2B). In the examination of variants presented in each mitochondrial haplogroup, we found that mutation count correlates with the haplogroups’ evolutionary lineage, which was also supported by earlier findings^16,28^ (Figure S2).

**Figure 2.**
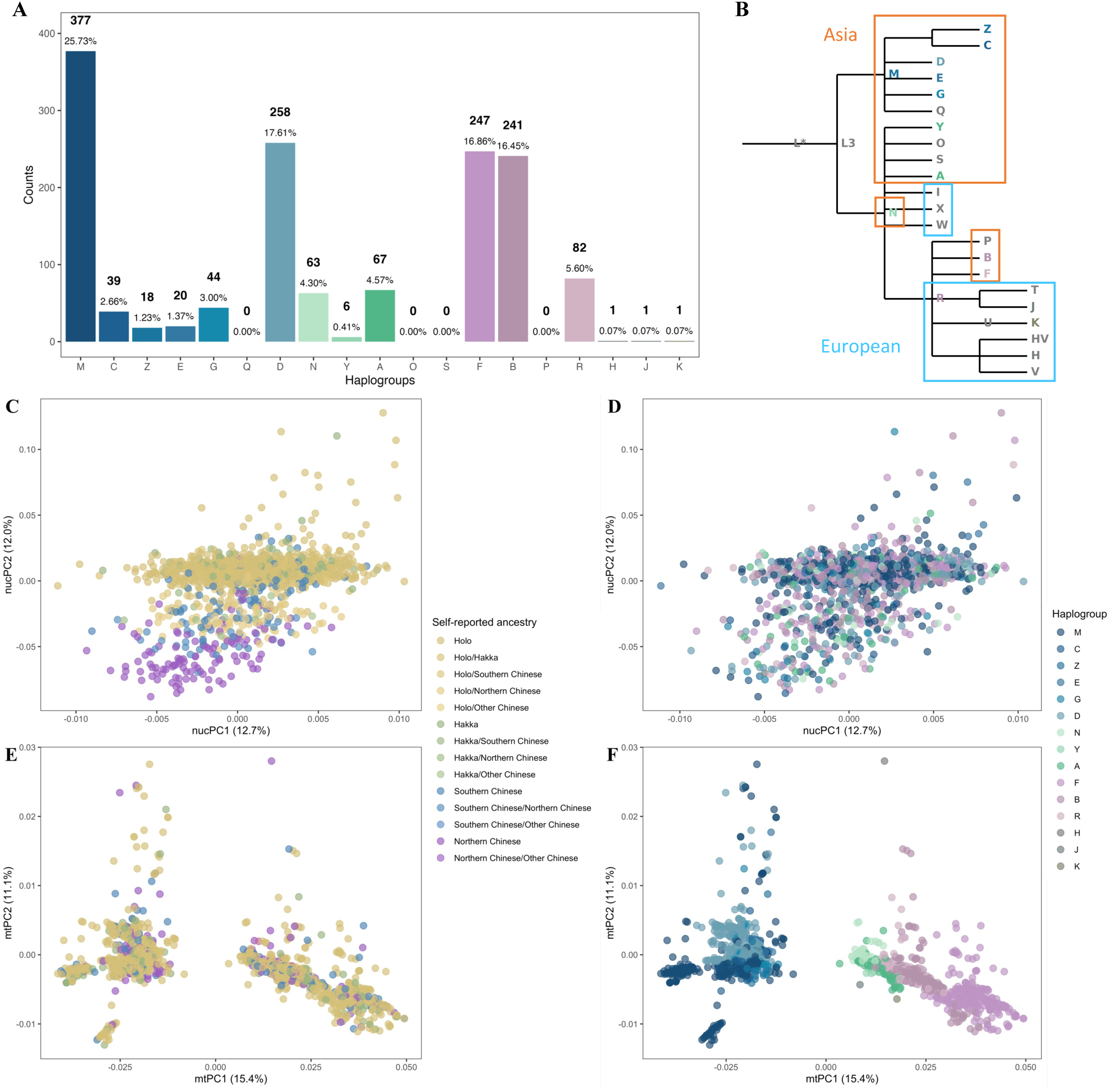
mtDNA haplogroups analysis in TWB. (A) The bar graph displays the prevalence of mtDNA haplogroups among TWB participants, arranged according to their relationships in the mitochondrial Phylotree. Haplogroups deriving from macrohaplogroup M are colored in shades of blue, those from N in shades of green, and those from R in shades of purple. For clarity, each haplogroup’s color is consistent across all subfigures. (B) A simplified tree depicting mtDNA haplogroups found among TWB participants, with each haplogroup uniquely colored. Asian and European haplogroups are delineated by distinct colors as defined in MITOMAP. Haplogroups not present in the TWB are shown in grey. (C and D) PCA analyses using nuclear variants from 1,492 participants. Participants are colored by self-reported parental ancestry in (C) and haplogroup assignment in (D). (E and F) PCA analyses using mtDNA variants from 1,492 participants. Participants are colored by self-reported maternal ancestry in (E) and haplogroup assignment in (F).

In contrast to the well-documented nuclear structure that reflects population structure within the Taiwanese population^14,15^, less is known about mitochondrial genome structure in Taiwan. Moreover, to appropriately control for population stratification—essential for robust association studies—it is necessary to elucidate the correlation between two genetic structures. We conducted principal component analyses (PCA) on both nuclear and mitochondrial genomes. Nuclear PCA revealed a high degree of homogeneity among the TWB population, with individuals reporting Northern Han Chinese ancestry positioned peripherally in the genetic landscape (Figure 2C). In contrast, the mitochondrial PCA revealed a clear clustering of individuals based on their haplogroup assignments (Figure 2F), reflecting anticipated diversity in mitochondrial genotypes. However, a limited correlation was observed between nuclear PCs and mitochondrial haplogroups, and no substantial correlation was evident between mitochondrial PCs and maternal ancestries (Figure 2D and 2E; Figure S3). These findings support the arguments made in the BBJ that there is no correlation between two genomes. Additionally, nuclear genome-derived PCs adequately account for the influence of population stratification when evaluating associations with mtDNA variants.

### Prevalence of confirmed pathogenic mtDNA variants in TWB

To comprehensively the mitochondrial genetic profile, we investigated the prevalence of 96 confirmed pathogenic mtDNA variants listed in MITOMAP across 1,465 WGS samples^19^. We identified 6 pathogenic variants with a VAF greater than 10%, indicating a carrier frequency of 0.556%, which aligns with earlier estimates of about 0.5%^16,28^. To extend our findings and enhance the scope of our study, we included 120,163 TWBv2 microarray samples. The TWBv2 SNP array was specifically designed to cover a wide range of disease-relevant variants and serves as a valued source for this purpose. This genotyping array includes 815 mtDNA variants, with 58 confirmed pathogenic variants.

From the microarray, we identified 20 pathogenic variants, revealing a carrier frequency of about 0.487%. In total, we pinpointed 23 confirmed pathogenic mtDNA variants within the TWB (Table S2). Many of these variants are associated with mitochondrial diseases known for their incomplete penetrance, including aminoglycoside-induced hearing loss, Leber’s Hereditary Optic Neuropathy (LHON) and mitochondrial encephalomyopathy, lactic acidosis, and stroke-like episodes (MELAS)^29^. Among the identified variants, m.1555A>G (rs267606617), linked to aminoglycoside-induced hearing loss, exhibited the highest frequencies in both WGS and microarray data. Other variants include the identification of LHON-associated variants m.4171A>G (rs28616230), m.11778G>A (rs199476112), and m.14484T>C and a MELAS-associated variant m.3697G>A (rs199476122). These findings contribute to the understanding of mitochondrial pathology within the Taiwanese population.

### Imputation of mtDNA variants

To explore the complex interplay between mtDNA variants and a range of complex traits, our study leveraged microarray data from the TWB. We imputed mtDNA variants from the microarray using a reference panel derived from the 1,465 WGS, enabling us to analyze mtDNA variants across 101,473 participants.

First, we rigorously validated the accuracy of genotyped mitochondrial variants in microarray by comparing AFs between shared mtDNA variants from the microarray and WGS data. We found strong concordance, except for one variant that showed a significant deviation, which was removed in the following analysis (Figure S4A). Next, we assessed the capability of the microarray data to detect low-level heteroplasmies. Utilizing a subset of 1,426 individuals with both WGS and microarray data, we evaluated whether genotype calls presented in WGS could be detected in microarray samples. Our results showed limitations of microarray in identifying heteroplasmic variants, particularly at heteroplasmy levels lower than 70% (Figure S4B). To address this, we excluded all heteroplasmies from the reference panel from the reference panel construction. Finally, to define the appropriate threshold for post-imputation quality control, we utilized the same subsets of 1,426 individuals and calculated the genotype concordance rate of imputed mtDNA variants within each VAF bin (Methods). Based on this analysis, we selected an INFO score >0.7 (estimated by IMPUTE2) to ensure high-quality imputed genotypes (Figure S4C). With the imputation quality ascertained, we refined our dataset through a workflow tailored for mtDNA variant imputation (Figure S5). After applying this quality filter and restricting AFs to within 0.1 deviation from WGS data, the final dataset included 563 high-confidence imputed mtDNA variants.

### Mitochondrial genome-wide association study

To comprehensively analyze the association between mtDNA variants and complex traits, we conducted mitochondrial genome-wide association analyses. We retained 306 mtDNA variants with an MAF greater than 0.01 for downstream analysis (Table S2). These variants demonstrated high allele frequency consistency with WGS data (Pearson r = 0.999).

Focusing on diseases prevalent among TWB participants, we included 48 binary traits for analysis (Table S3). Additionally, we leveraged 38 quantitative traits derived from blood biomarkers, excluding outliers and applying rank inverse-normal transformations as appropriate (Table S4; Methods). Using the imputed dosages (0 or 2) of the individuals, we conducted regression analyses with adjustment for sex, age, age and sex interaction, genotype batches, and potential population stratification by including the top 10 nucPCs as covariates (Figure S6).

Despite no mtDNA variants meeting the conservative significance threshold, several variants were observed to associate with traits at a lenient threshold adjusting for total mtDNA variant counts (P = 0.05/306 = 1.4 × 10^−4^) (Figure 3; Table S5 and S6).

**Figure 3.**
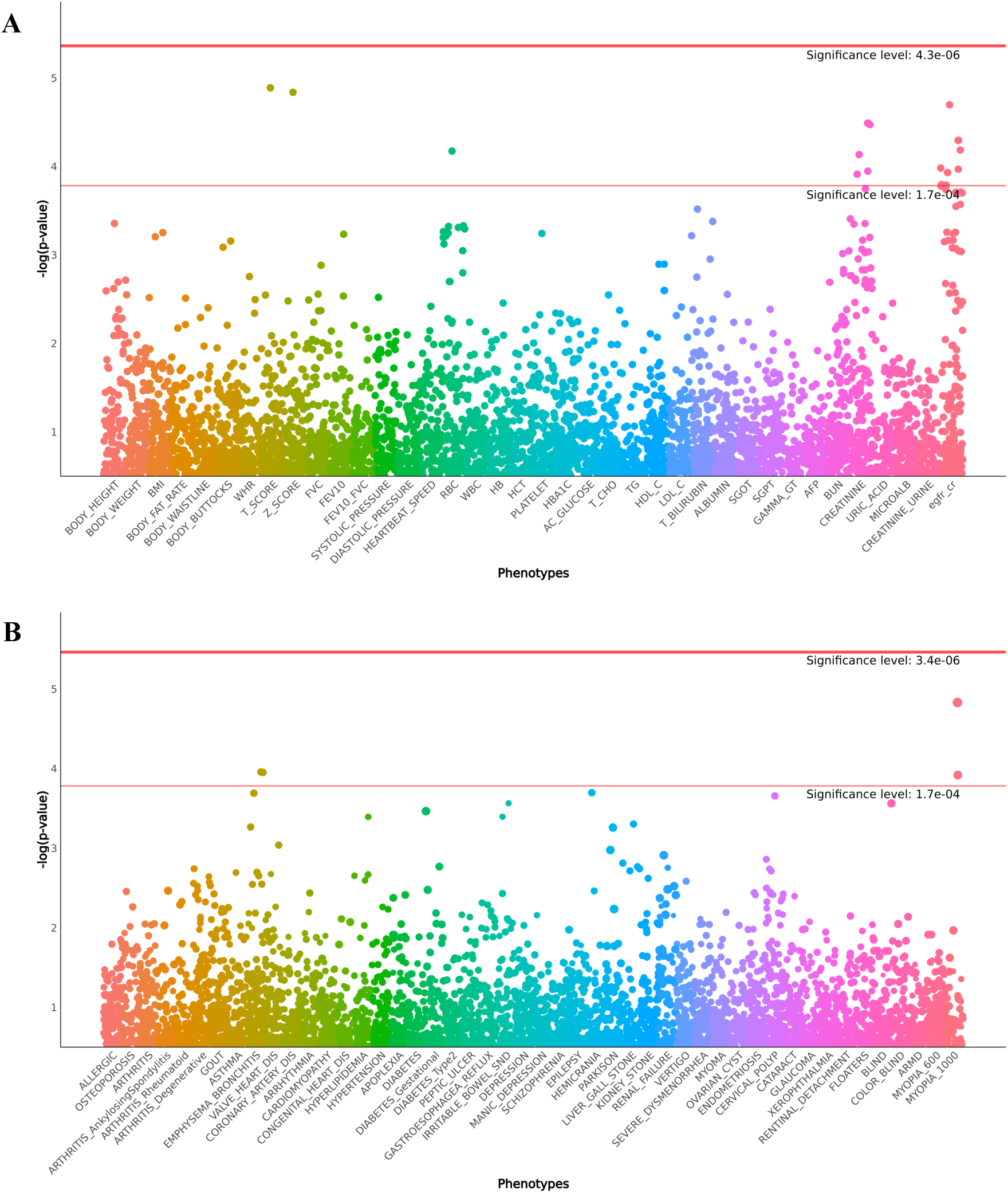
Manhattan plots of mtDNA variant associations with complex traits. The Manhattan plots show the association results of mtDNA variants with multiple complex traits analyzed using a generalized linear model (GLM). Each dot represents a mtDNA variant analyzed for association with a specific phenotype, sorted by position along the x-axis. The colors represent different phenotypes, each denoted by a unique color. Association analyses for quantitative and binary traits are shown in a and b, respectively. Two significance thresholds are shown. The lower red line represents a lenient threshold adjusted by the number of variants (P = 1.4 × 10^−4^), while the upper orange line represents a stricter threshold adjusted further by the number of phenotypes.

### Association of mtDNA variants with high myopia

In our study, we identified associations between two mtDNA variants and high myopia. The variants rs28570593 (m.5054 A>G) and rs367778601 (m.5147A>G) exhibited odds ratios (ORs) of 2.47 (P = 1.63 × 10^−5^) and 1.96 ( P = 1.08 × 10^−4^), respectively, suggesting their potential links to the condition. Both variants are synonymous variants located on the *MT-ND2* gene, with the mere linkage between them (*r*^2^=0.148) indicating they may independently contribute to high myopia (Figure 4). The first variant, rs28570593, showed a higher prevalence in cases (0.056) compared to controls (0.010). The second variant, rs367778601, also displayed a significant case-control frequency difference, with allele frequencies of 0.080 in cases versus 0.023 in controls. These findings are consistent with previous research that reported associations of different variants on the MT-ND2 gene with high myopia in the Han Chinese population.^30^.

**Figure 4.**
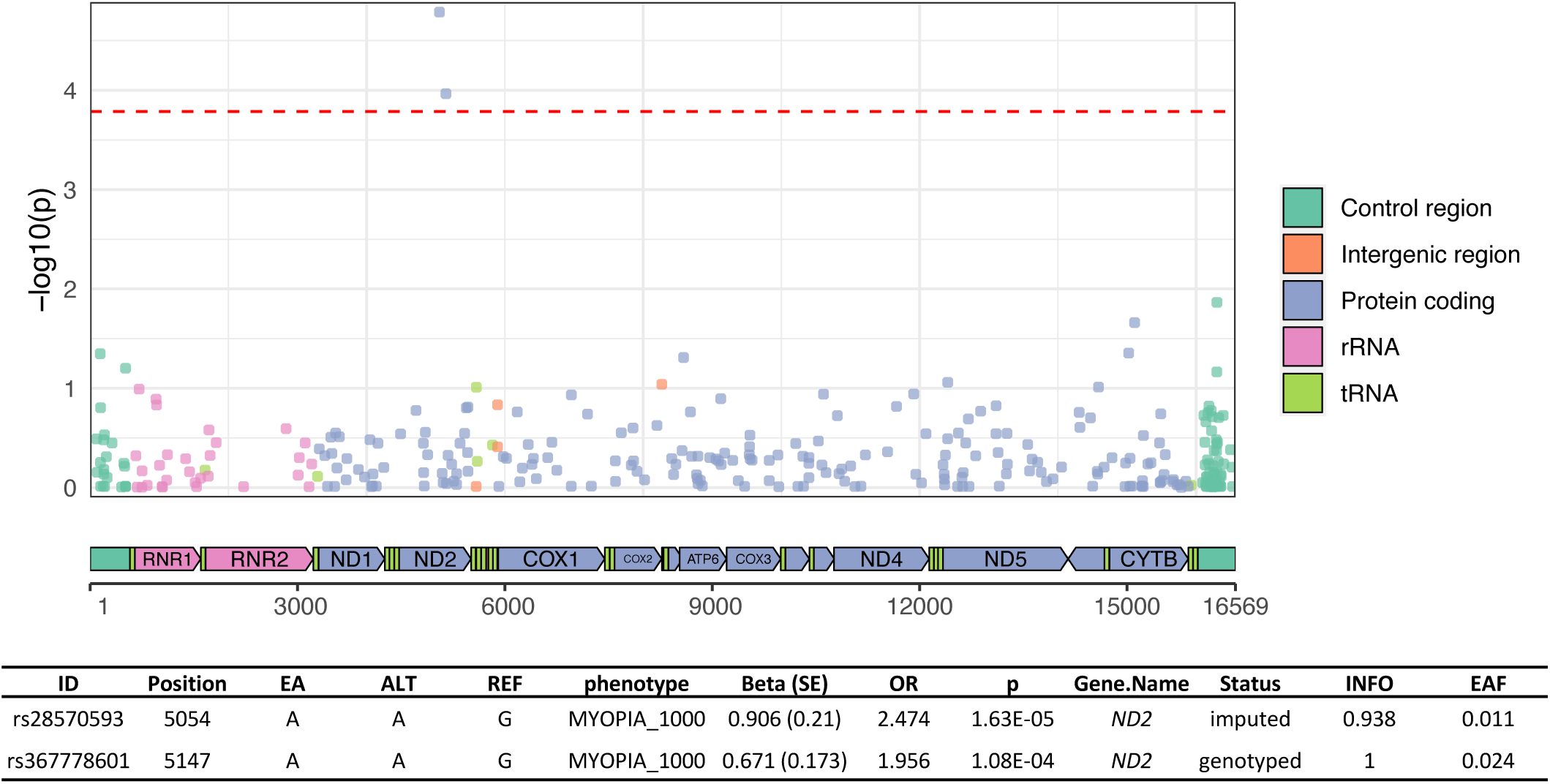
Manhattan plot of mtDNA variants associated with high myopia. The plot illustrates the –log10(p-values) of mtDNA variants analyzed for their association with high myopia, sorted by their position on the mitochondrial genome. Variants are color-coded based on their genomic location: control region (green), intergenic region (orange), protein-coding region (purple), rRNA genes (pink), and tRNA genes (light green). The red dashed line indicates the lenient significance threshold.

### Association of mtDNA variants with renal function biomarkers

We identified 14 mtDNA variants associated with biomarkers indicating renal function. specifically serum creatinine (Scr) levels and estimated glomerular filtration rate (eGFR) (Figure 5A; Table 1). Due to the lack of recombination in the mitochondrial genome^31^, we hypothesized that these variants may share common haplotypes. To investigate this, we first calculated pairwise linkage disequilibrium (LD) *r*^2^ focused on common mtDNA variants (MAF >0.05) using PLINK^21^. The results indicated multiple common haplotypes spanning the entire genome (Figure S7).

**Figure 5.**
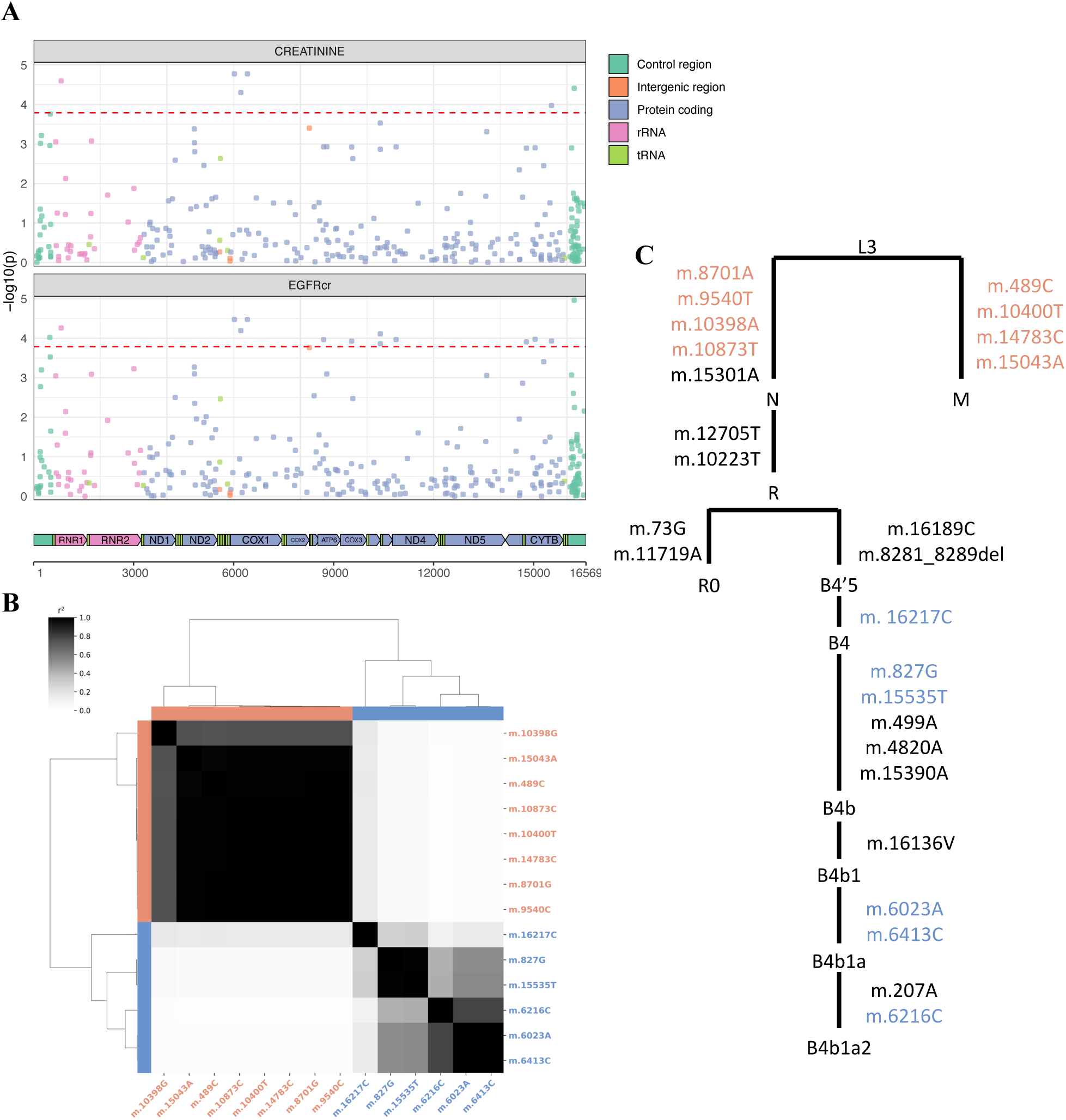
mtDNA variants associated with renal function and phylogenetic lineages. (A) The Manhattan plot illustrates mtDNA variants assessed for their association with renal function biomarkers: Scr levels and eGFR. Variants are color-coded based on their genomic location: control region (green), intergenic region (orange), protein-coding region (purple), rRNA genes (pink), and tRNA genes (light green). The red dashed line indicates the lenient significance threshold. (B) The heatmap displays the pairwise LD *r*² among the identified mtDNA variants, clustered to highlight two main association sets (orange and blue). LD values were calculated using PLINK.

**Table 1.** Renal function-associated mtDNA variants in the Taiwanese population.

| Phenotype | POS | REF | ALT | Gene (Consequence) | p | Beta (SE) | Status | INFO | Freq. | Haplogroup |
| --- | --- | --- | --- | --- | --- | --- | --- | --- | --- | --- |
| Scr | 827 | A | G | *MT-RNR1*<br>(non_coding_transcript_exon_variant) | 2.54E-05 | -0.023<br>(0.006) | genotyped | 1 | 0.044 | B4b |
| Scr | 6023 | G | A | *MT-COI*<br>(synonymous_variant) | 1.68E-05 | -0.032<br>(0.007) | imputed | 0.982 | 0.024 | B4b1a |
| Scr | 6216 | T | C | *MT-COI*<br>(synonymous_variant) | 4.99E-05 | -0.029<br>(0.007) | genotyped | 1 | 0.025 | B4b1a2 |
| Scr | 6413 | T | C | *MT-COI*<br>(synonymous_variant) | 1.68E-05 | -0.032<br>(0.007) | imputed | 0.995 | 0.024 | B4b1a |
| Scr | 15535 | C | T | *MT-CYB*<br>(synonymous_variant) | 1.07E-04 | -0.021<br>(0.006) | genotyped | 1 | 0.044 | B4b |
| Scr | 16217 | T | C | D-LOOP<br>(intergenic_variant) | 3.89E-05 | -0.014<br>(0.003) | imputed | 0.941 | 0.135 | B4 |
| eGFRcr | 489 | T | C | D-LOOP<br>(intergenic_variant) | 9.53E-05 | -0.008<br>(0.002) | imputed | 0.971 | 0.51 | M |
| eGFRcr | 827 | A | G | *MT-RNR1*<br>(non_coding_transcript_exon_variant) | 5.52E-05 | 0.021<br>(0.005) | genotyped | 1 | 0.044 | B4b |
| eGFRcr | 6023 | G | A | *MT-COI*<br>(synonymous_variant) | 3.37E-05 | 0.029<br>(0.007) | imputed | 0.982 | 0.024 | B4b1a |
| eGFRcr | 6216 | T | C | *MT-COI*<br>(synonymous_variant) | 6.43E-05 | 0.027<br>(0.007) | genotyped | 1 | 0.025 | B4b1a2 |
| eGFRcr | 6413 | T | C | *MT-COI*<br>(synonymous_variant) | 3.37E-05 | 0.029<br>(0.007) | imputed | 0.995 | 0.024 | B4b1a |
| eGFRcr | 8701 | A | G | *MT-ATP6*<br>(missense_variant) | 1.09E-04 | -0.008<br>(0.002) | imputed | 0.995 | 0.508 | N (m.8701A) |
| eGFRcr | 9540 | T | C | *MT-CO3*<br>(synonymous_variant) | 1.19E-04 | -0.008<br>(0.002) | imputed | 0.998 | 0.508 | N (m.9540T) |
| eGFRcr | 10398 | A | G | *MT-ND3*<br>(missense_variant) | 7.77E-05 | -0.008<br>(0.002) | genotyped | 1 | 0.573 | N<br>(m.10398A) |
| eGFRcr | 10400 | C | T | *MT-ND3*<br>(synonymous_variant) | 1.39E-04 | -0.008<br>(0.002) | genotyped | 1 | 0.508 | M |
| eGFRcr | 10873 | T | C | *MT-ND4*<br>(synonymous_variant) | 1.09E-04 | -0.008<br>(0.002) | genotyped | 1 | 0.505 | N<br>(m.10873T) |
| eGFRcr | 14783 | T | C | *MT-CYB*<br>(synonymous_variant) | 1.24E-04 | -0.008<br>(0.002) | imputed | 0.996 | 0.509 | M |
| eGFRcr | 15043 | G | A | *MT-CYB*<br>(synonymous_variant) | 1.06E-04 | -0.008<br>(0.002) | genotyped | 1 | 0.509 | M |
| eGFRcr | 15535 | C | T | *MT-CYB*<br>(synonymous_variant) | 1.18E-04 | 0.02<br>(0.005) | genotyped | 1 | 0.044 | B4b |
| eGFRcr | 16217 | T | C | D-LOOP<br>(intergenic_variant) | 1.09E-05 | 0.014<br>(0.003) | imputed | 0.941 | 0.135 | B4 |

The table presents renal function-associated variants, detailing their positions in the reference genome NC\_012920.1, reference (REF) and alternative (ALT) alleles, gene location, variant consequences, and defining haplogroups. Statistical results are presented with p-values, effect sizes (Beta), and standard errors (SE) of the association between the ALT allele and the phenotype. Additional information includes imputation status, IMPUTE2 information scores (INFO), and allele frequencies from genotyping arrays in the TWB.

To further elucidate whether mtDNA variants associated with renal function belong to the same haplotypes, we categorized these variants based on their LD *r*^2^, revealing two distinct clusters (Figure 5B). The first set primarily comprises ancestral markers for the divergent mitochondrial super-haplogroups M and N. Variants defining super-haplogroup M are associated with increased risk factors for impaired renal function, including higher Scr levels and lower eGFR. Conversely, alleles commonly found in super-haplogroup N correlate with decreased Scr levels and higher eGFR, suggesting a potentially protective effect on kidney function. The second set of variants belongs to haplogroup B4b, a subgroup within the N lineage. These B4b variants demonstrate associations with lower Scr levels and higher eGFR, further supporting a protective effect against renal impairment.

### B4b haplogroup is associated with renal function

Our results suggest that a specific haplogroup background can affect susceptibility to renal function. To investigate this further, we conducted an analysis of haplogroup-specific effects on renal function markers. We utilized imputed and quality-controlled mtDNA variants to assign haplogroups to individuals using HaploGrep2^17^, achieving a mean quality score of 0.906 (Figure S8). We then tested the association between sub-haplogroups and renal function markers, specifically Scr and eGFR. Sub-haplogroups were included in the analysis if they were carried by more than 500 individuals.

Our analysis revealed that the B4b sub-haplogroup was significantly associated with decreased Scr (P = 8.15 × 10^−5^, β (SE) = −0.06 (0.01)) and increased eGFR (P = 7.33 × 10^−5^, β (SE) = 0.05 (0.01)) (Figure S9; Table S7 and S8). These findings suggest a potentially protective role of the B4b haplogroup in renal function. However, when we conducted an association analysis between the B4b haplogroup and self-reported renal failure status, no significant association was observed. This discrepancy suggests that while B4b may have a beneficial effect on renal function markers, its influence may be subtle and not directly translate to a reduced incidence of clinically diagnosed renal failure.

## Discussion

In this study, we comprehensively analyzed the mtDNA of individuals from the TWB. Our findings provide valuable insights into the genetic characteristics of the Taiwanese population and contribute to our understanding of mitochondrial genetics and its implications for health and disease.

Our mtDNA haplogroup analysis revealed a predominance of Asian-associated haplogroups (M, D, F, and B), aligning with the expected genetic heritage of the Taiwanese population. However, the distinct patterns revealed by mtDNA-based PCA compared to autosomal PCA suggest limitations in using mtDNA alone to capture ancestral variations within the TWB. Our findings indicate no direct correlation between nuclear and mitochondrial genomes, supporting similar observations from the BBJ study^12^. This contrasts with findings from the UKB, where associations between mitochondrial sub-haplogroups and nuclear PCs were identified^13^. This discrepancy highlights the complexity of genetic correlation and underscores the need for a more integrative approach to elucidate the full spectrum of associations between the two genomes in both genotype-and population-level in diverse populations.

We conducted a large-scale mitochondrial-genome-wide association study across 86 traits and 306 mtDNA variants in 101,473 Taiwanese participants. Our study demonstrated a successful imputation using WGS as a reference panel and revealed abundant mtDNA variants risk on complex traits. We identified associations between mtDNA variants and high myopia (HM). Specifically, two variants in the *MT-ND2* gene—rs28570593 and rs367778601—were linked to HM, particularly in individuals requiring spherical corrections of –10 diopters (D) or more. The *MT-ND2* gene encodes NAHD dehydrogenase 2, a component of complex I in the mitochondrial respiratory chain system, crucial for cellular energy production^32^. Previous studies have indicated *MT-ND2* variants, such as m.5244G>A^33^ and m.4640C>A^34^, in Leber hereditary optic neuropathy (LHON), suggesting a role of *MT-ND2* dysfunction in ocular diseases. In a recent study, Xing et al. identified nine novel mitochondrial variants associated with HM, including rs370378529 in *MT-ND2,* with an odds ratio (OR) of 5.25^30^. These findings underscore the pivotal role of complex I in cellular energy metabolism, where subtle dysfunctions may not cause overt disease but contribute to conditions like myopia.

mtDNA variants have emerged as significant factors in kidney function^35^, reflecting the high mitochondrial content and oxygen demand of renal tissues^35^. In this study, we identified 14 mtDNA variants associated with renal function biomarkers, highlighting the potential relationship between mtDNA and kidney health. These mtDNA variants, while seemingly unrelated and distributed across the whole mitochondrial genome, can be categorized into two distinct sets: ancestral variants linked to the macrohaplogroup M diverges from L3, as well as the B4b sub-haplogroup. Notably, ancestral variants for macrohaplogroup M were associated with worse renal function indicators, with the missense variant m.10398A>G in the *MT-ND3* gene showing the most significant association with decreased eGFR. The G allele of this variant and the association of risk trends in renal function are reported in previous studies^36^. This variant is associated with impaired mitochondrial function, specifically affecting the function of Complex I of the electron transport chain, which is critical for energy-intensive processes in renal cells^32^.

Conversely, the B4b sub-haplogroup was significantly associated with improved renal function markers, including decreased Scr and increased eGFR. While the haplogroup is predominantly found in East and Southeast Asia^37^, the finding underscores the value of conducting association studies in non-European cohorts to uncover population-specific genetic influences on renal function.

Comparing our findings with other large-scale studies, such as the UK Biobank and Japan Biobank^12,13,36^, reveals both consistencies and population-specific differences in mtDNA variants’ implications in renal function indicators. Most variants associated with serum renal indicators in these studies were either absent or not replicated in our TWB cohort, likely reflecting population-specific genetic influences on renal function. However, the variant m.3010G>A, located on the *MT-RNR2* gene, showed a consistent association with decreased renal function across studies, being linked to increased cystatin C, decreased eGFR^cy^, and decreased eGFR^crcy^ in the UK Biobank and decreased eGFR in our cohort. This consistency across diverse populations strengthens the evidence for this variant’s role in kidney function.

A recent study from the INTERVAL dataset demonstrated that common mtDNA variants influence blood *N*-formylmethionine (fMet) levels, a critical amino acid in mitochondrial translation. Variants in mtDNA haplogroups Uk and H4 were linked to elevated fMet levels, potentially disrupting mitochondrial protein synthesis and degradation. This disruption could impair kidney function and contribute to age-related diseases by altering energy metabolism and protein dynamics^38^. Notably, the m.10398A>G variant in our cohort, associated with decreased eGFR, also presented in the INTERVAL dataset and linked to increased fMet levels, providing a potential mechanistic explanation for its effect on renal function.

Despite its contribution, our study still has limitations. First, while we comprehensively dissected the mitochondrial genetic profiles in the Taiwanese population, our analysis predominantly identified SNVs and indels, but larger structural variants (SVs), which could be equally significant, remain undetected. Second, due to the constrained sample size in the association analyses, our focus was primarily on microarray data, thus limiting to homoplasmic or near-homoplasmic variants. This may overlook the role of heteroplasmic variants, which are crucial in certain diseases^39^. Moreover, our analysis relies on self-reported data from questionnaire surveys, which may introduce inaccuracies due to biases or errors in self-reporting. Finally, given that some mtDNA variants were not imputed due to the limited genotyped SNPs and the nature of common haplotypes spanning the whole mitochondrial genome, it is challenging to map the causal variants. While the study provides insights into the association between mtDNA variants and complex traits, further replication datasets or molecular studies are necessary to elucidate the mechanisms influencing disease processes.

Our analysis revealed that neither the mtDNA variants nor haplogroups we identified were associated with self-reported renal failure status. This observation suggests that the effects of these variants may be subtle and act as phenotype-modifiers^40^, with their cumulative impact potentially increasing the risk of poor kidney outcomes over the long term. Thus, understanding the role of mitochondrial genetic background is crucial for gaining insights into phenotypic variability.

In conclusion, our study provides a comprehensive characterization of mtDNA within the TWB, significantly enhancing our understanding of mtDNA diversity and its implications for health and disease in the Taiwanese population. These findings emphasize the critical importance of including diverse genetic backgrounds in mitochondrial research by revealing insights into genetic diversity and the pivotal roles of mtDNA variants in complex traits.

## Data Availability

All data produced in the present study are available upon reasonable request to the authors

## Figures

**Figure S1.**
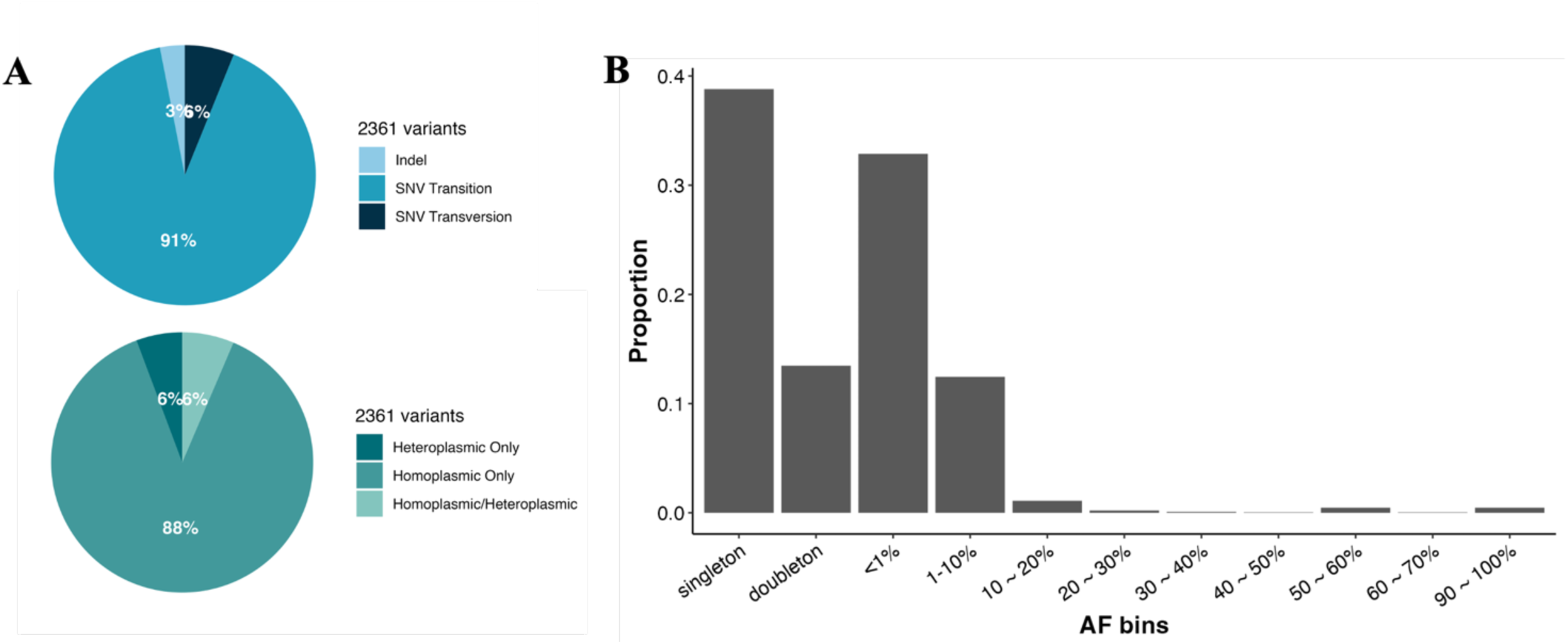
Distribution and classification of mtDNA variants in WGS analyses. (A) The left pie chart depicts the proportion of variants by different variants. The right pie chart shows the classification of variants that are homoplasmic-only, heteroplasmic-only, or occurring in both types. (B) The bar graph of AF bins for mtDNA.

**Figure S2.**
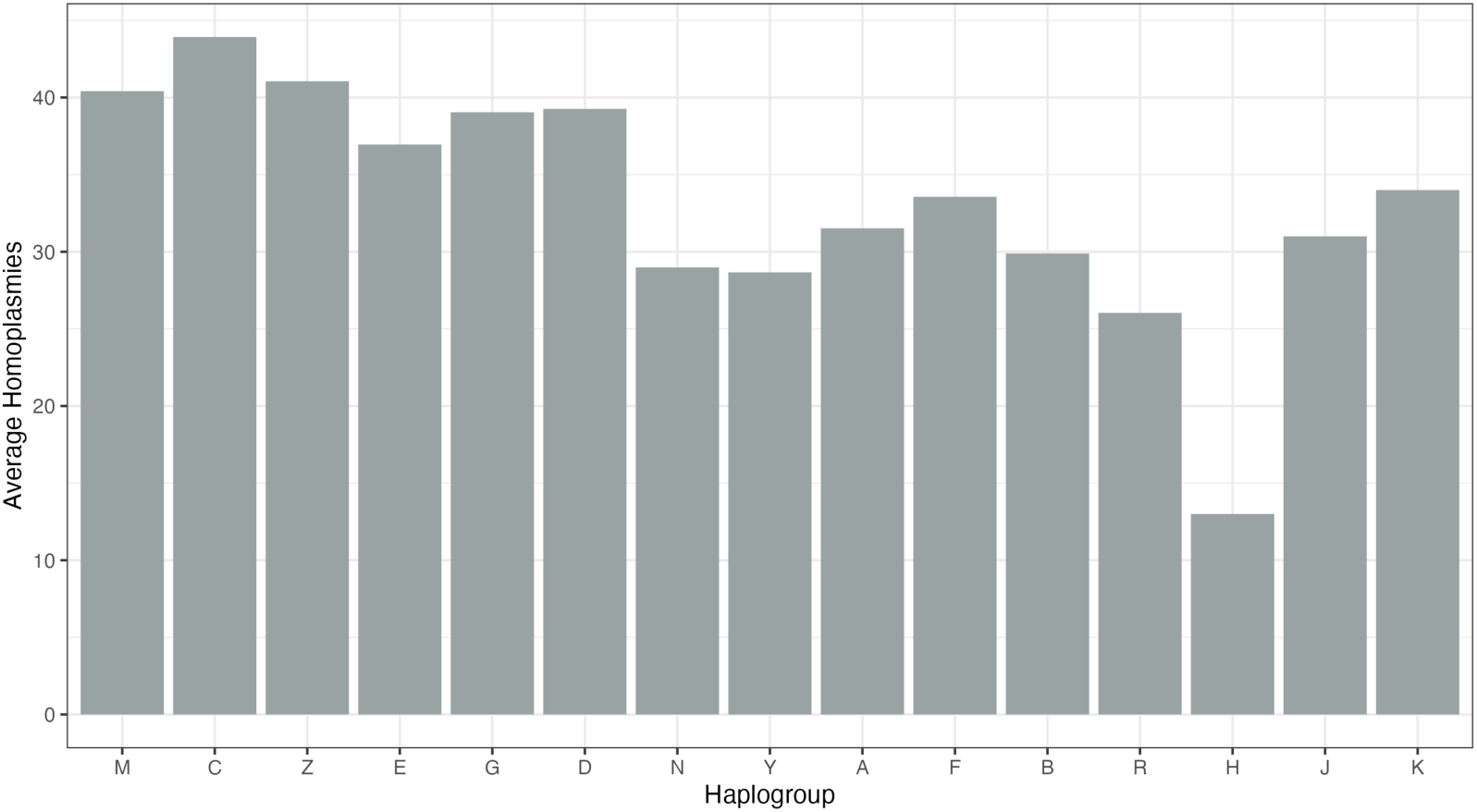
Average homoplasmies within haplogroups. The bar graph displays the average number of mtDNA homoplasmic variants individuals carry within each haplogroup. The order of haplogroup was aligned with Figure 2A.

**Figure S3.**
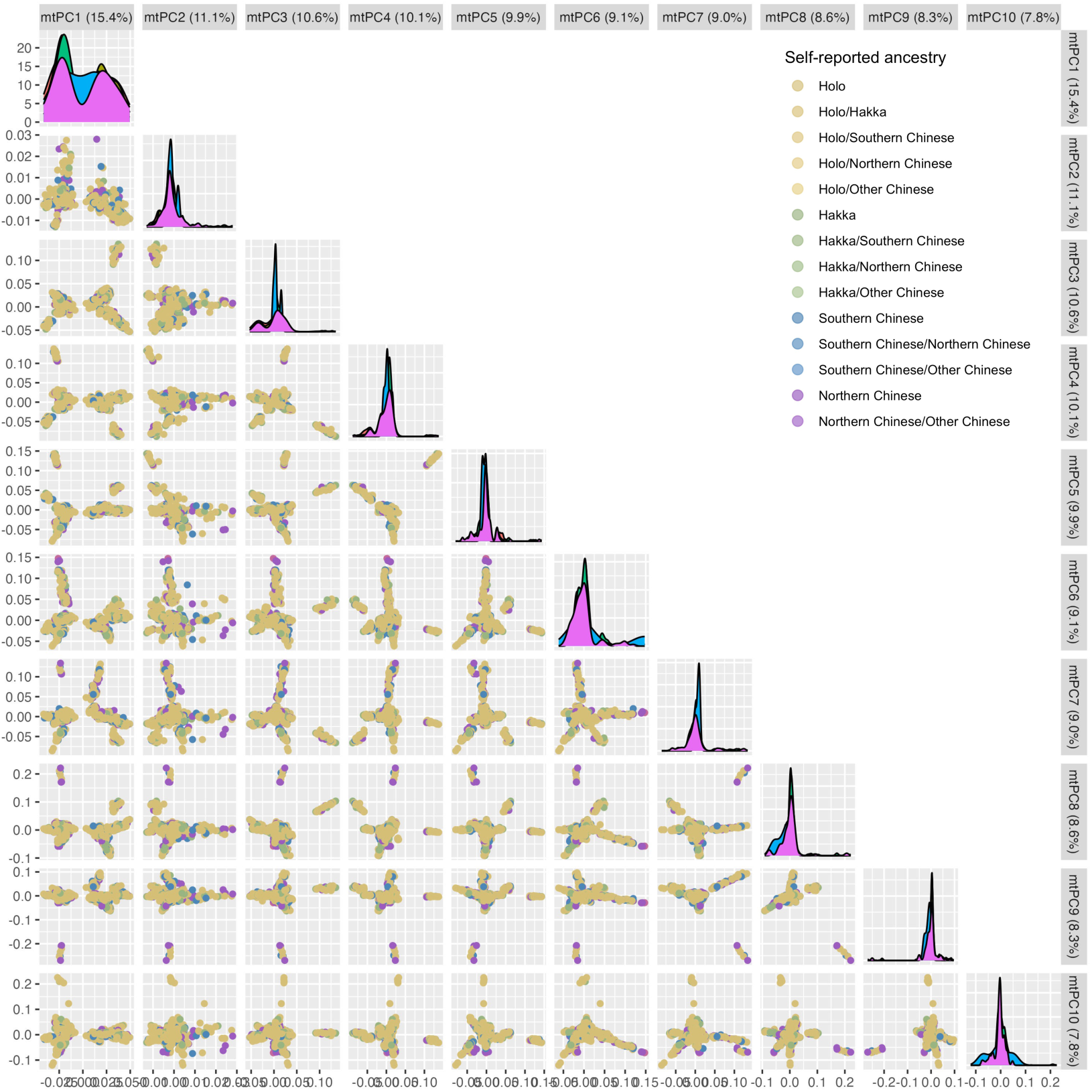
Pairwise plots of mitochondrial PCs (mtPC1 to mtPC10). Participants are colored according to their self-reported maternal ancestry, with density plots shown along the diagonal.

**Figure S4.**
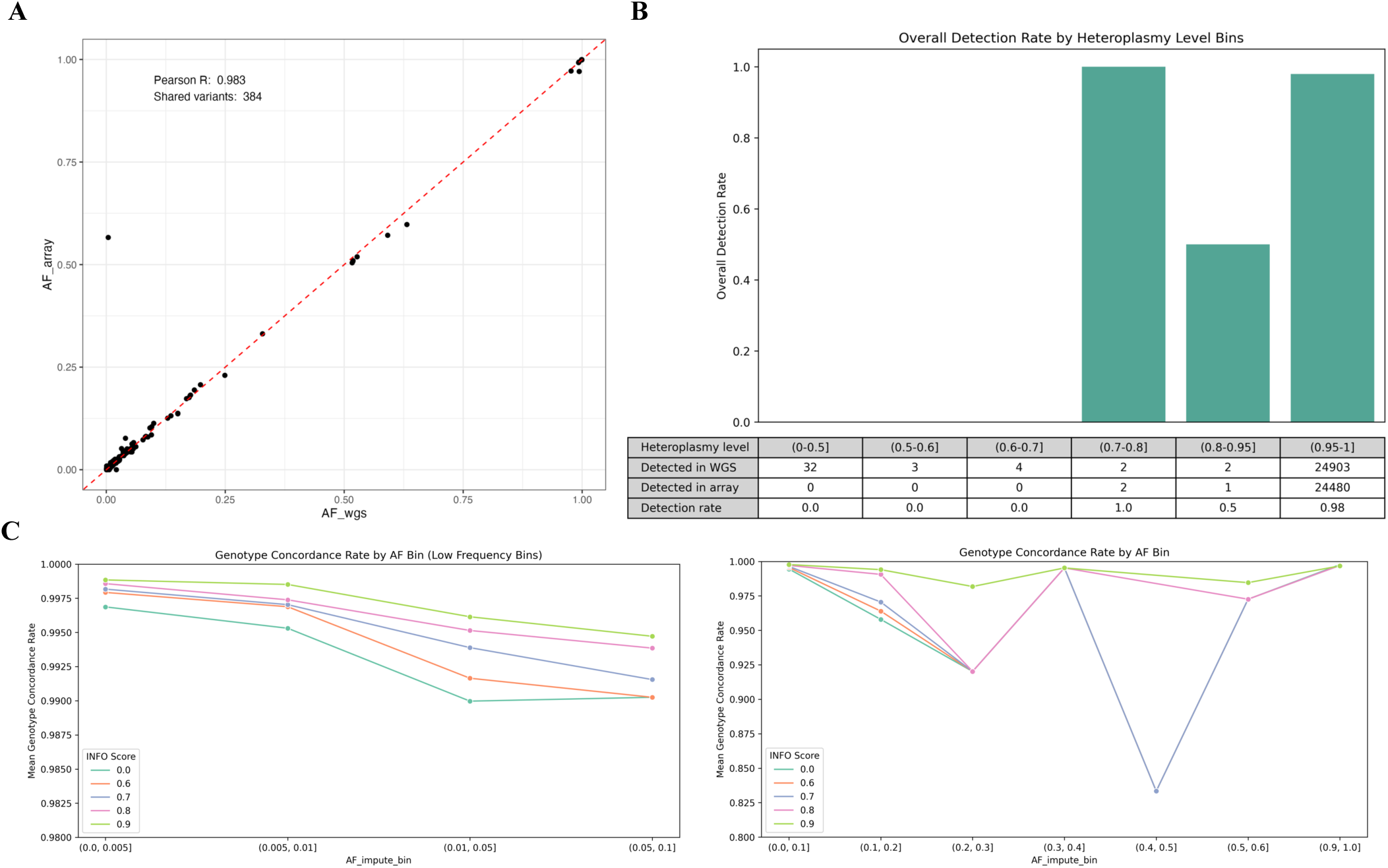
Evaluation of mtDNA variants imputation. (A) The plot illustrates the comparison of AFs between microarray and WGS data. (B) The bar graph displays the overall detection rates of mtDNA variants by heteroplasmy levels. Below the graph, a table details the number of genotype calls detected in both WGS and the microarray, along with the microarray’s detection rate. (C) This plot demonstrates the genotype concordance rates across various AF bins at different INFO score thresholds, with separate analyses for rare and common variants presented in the left and right panels, respectively.

**Figure S5.**
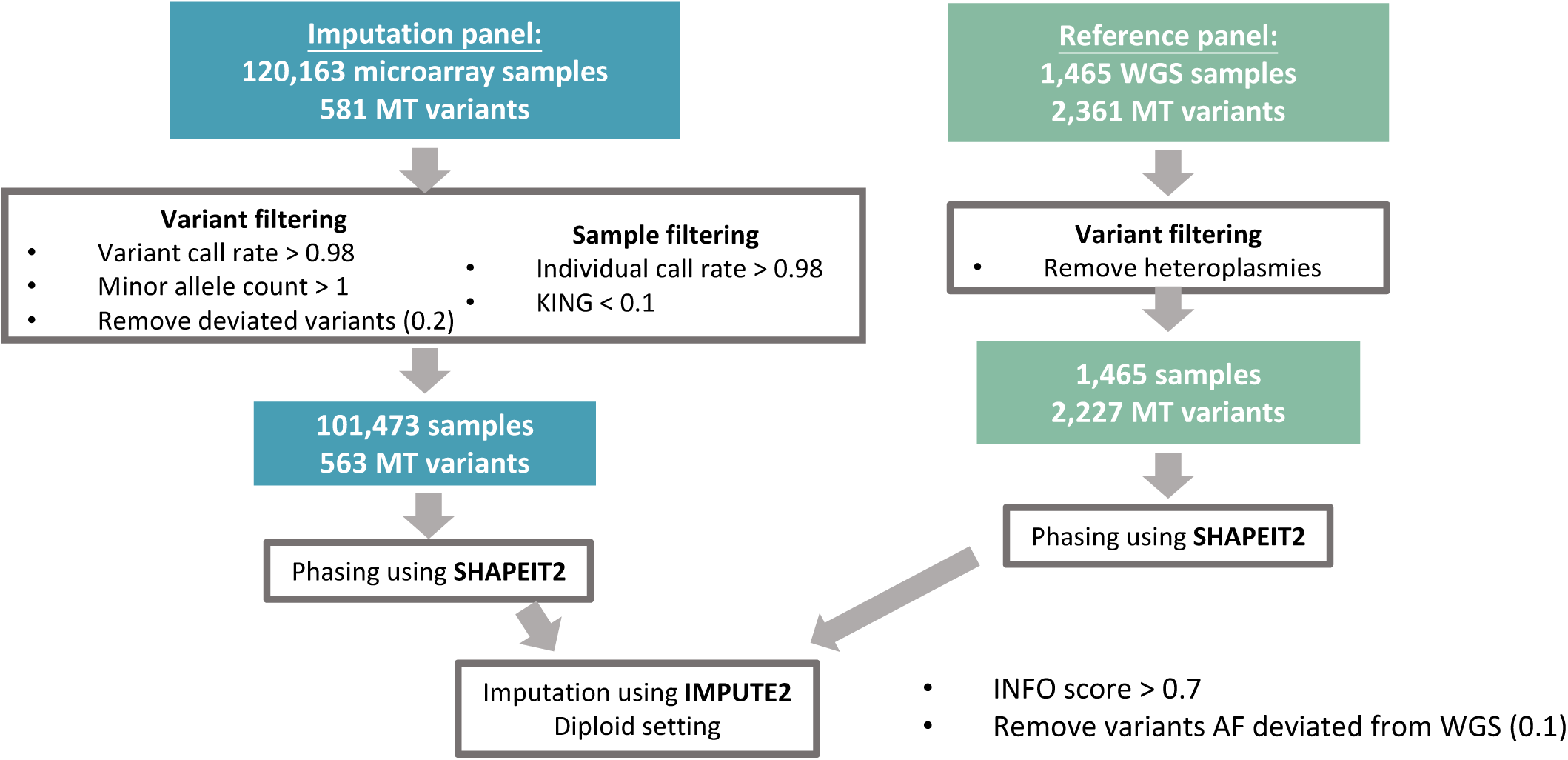
Overview of the mtDNA variant imputation process. The flowchart outlines the imputation workflow, including participant and variant counts, filtering criteria, and software (bold) used throughout the analysis.

**Figure S6.**
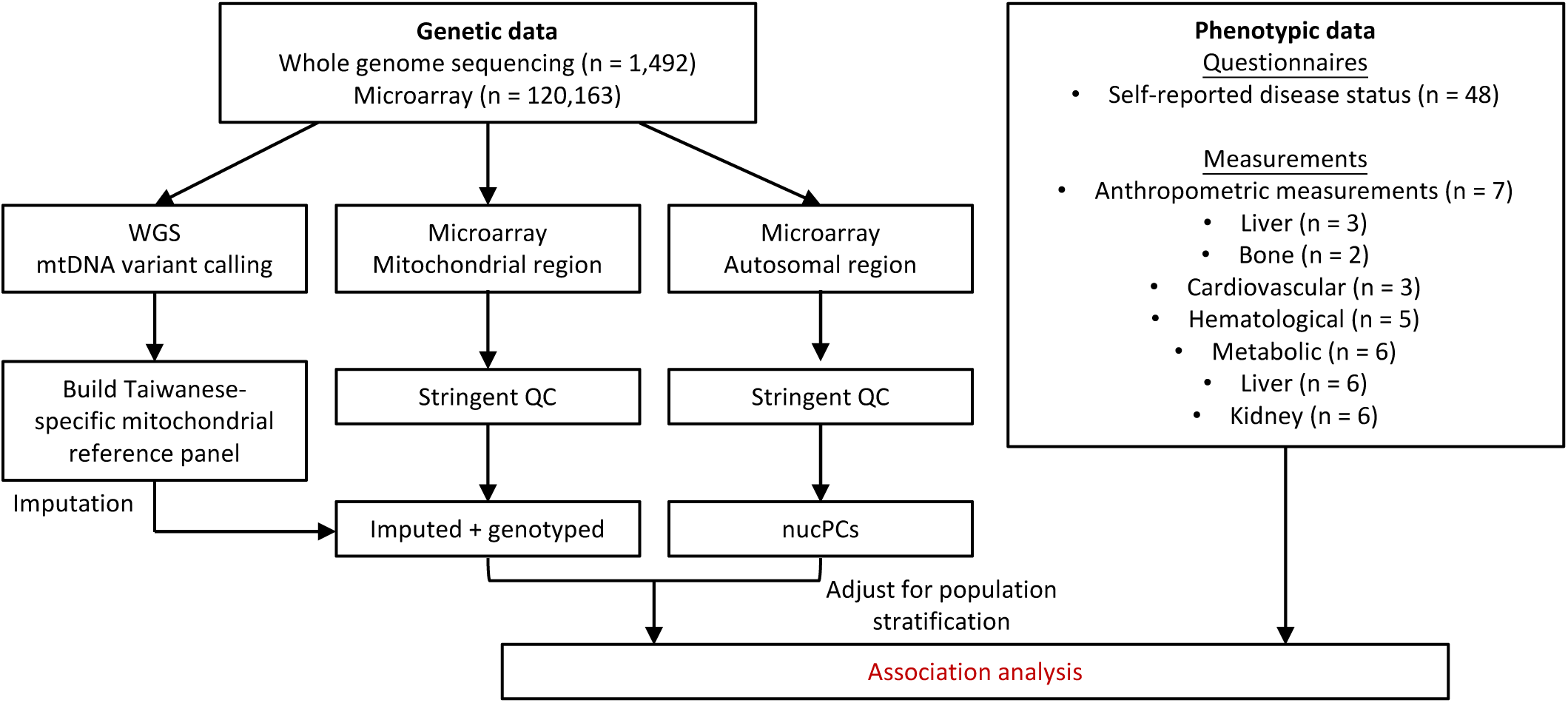
Mitochondrial genome-wide association analysis workflow. The schematic outlines the workflow for mtDNA variant association analysis using genetic data from 1,492 WGS and 120,163 microarray samples. This includes mtDNA variant calling, quality control, and imputation, followed by association analyses with phenotypic data from questionnaires and measurements relevant to complex traits.

**Figure S7.**
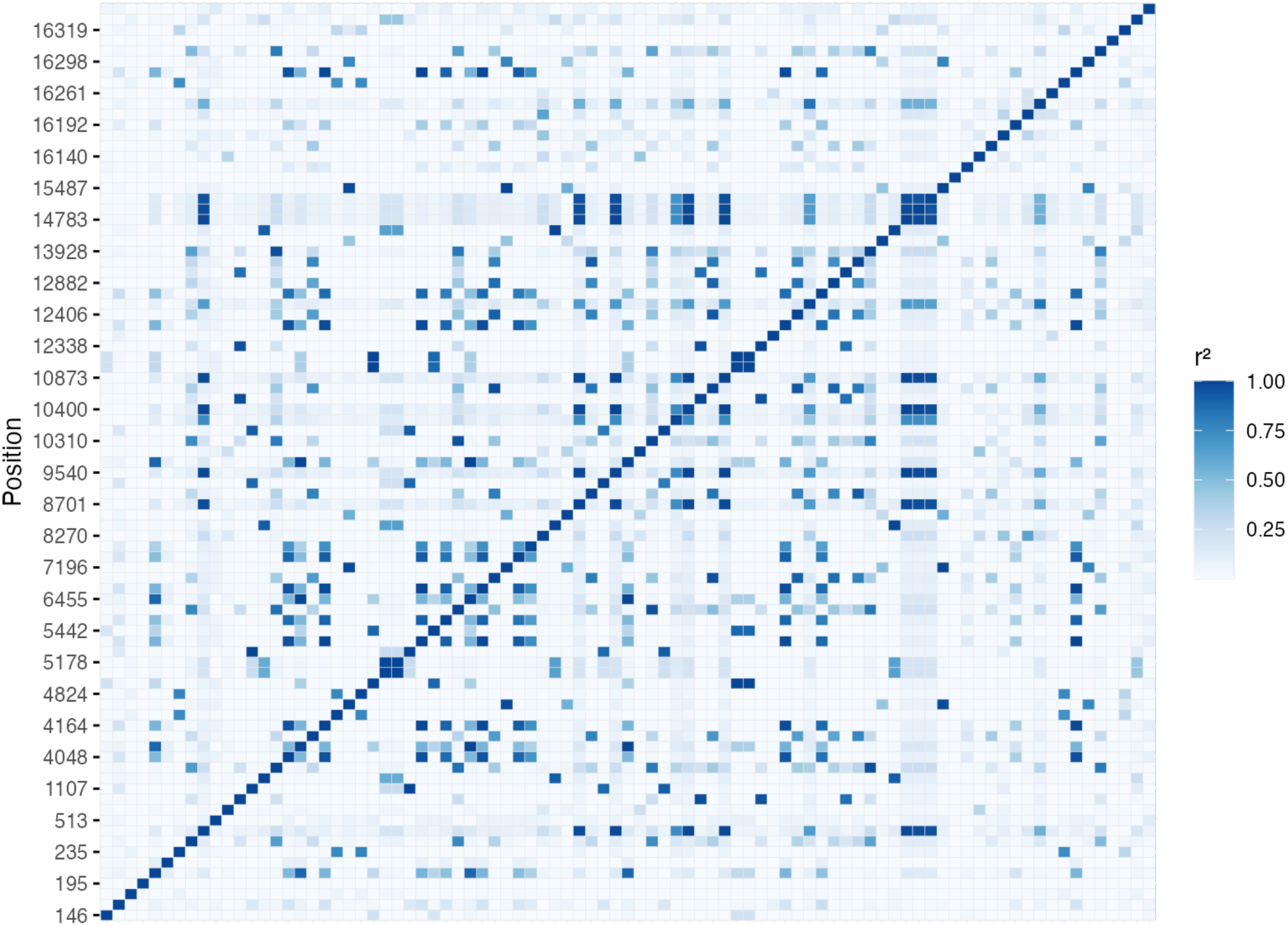
Pairwise LD matrix of mtDNA variants. The heatmap illustrates the pairwise LD patterns across homoplasmic mitochondrial DNA variants with an AF ≥0.05. Each cell represents the *r*² value between pairs of mtDNA variants, indicating the level of correlation. Darker shades indicate higher *r*^2^.

**Figure S8.**
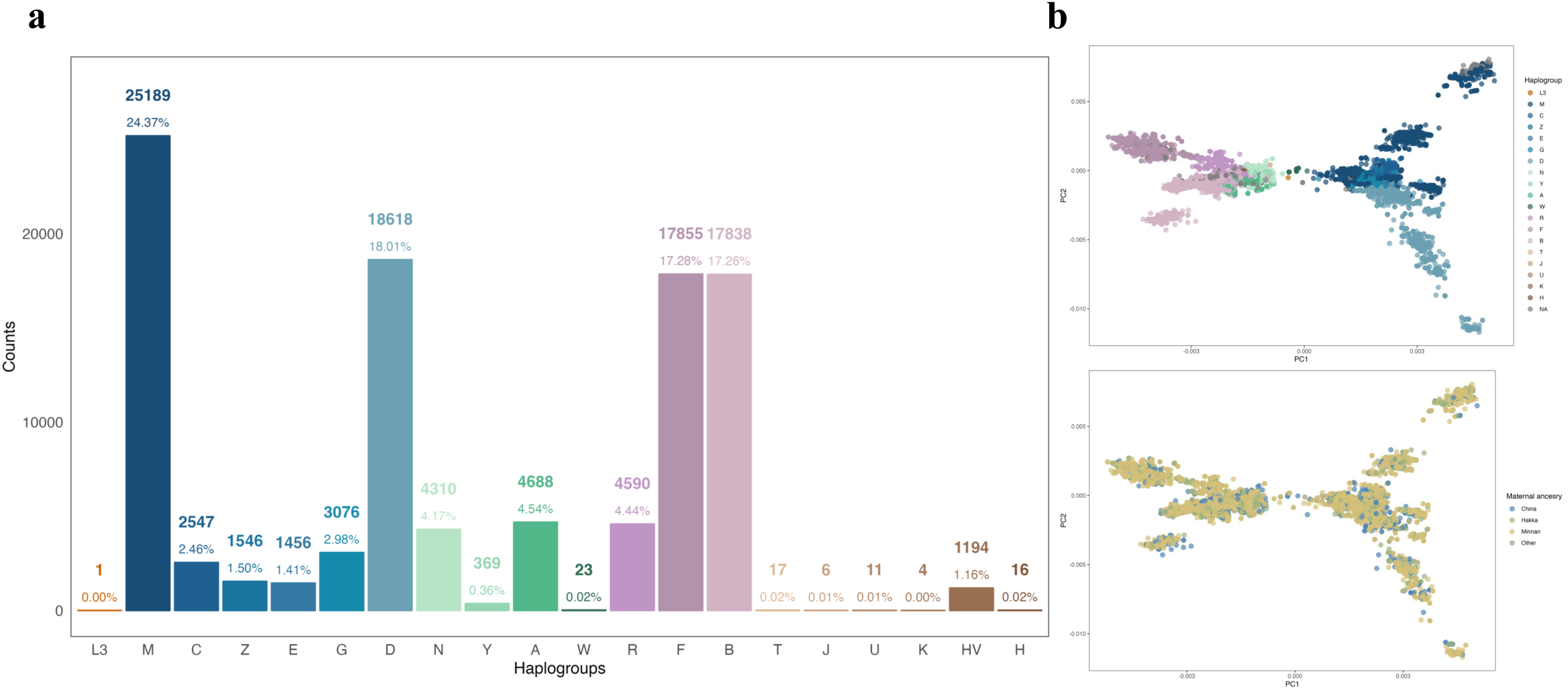
mtDNA haplogroups analysis in TWB using genotyping array. (A) The bar graph shows the prevalence of mtDNA haplogroups among TWB participants, with assignments based on microarray genotypes. The haplogroups are organized according to their relationships in the mitochondrial Phylotree. Haplogroups deriving from macrohaplogroup M are colored in shades of blue, those from N in shades of green, and those from R in shades of purple and brown. (B and C) PCA analyses of mtDNA variants derived from the genotyping array. The upper plot illustrates individuals colored by their haplogroup assignment, and the lower plot is colored by self-reported maternal ancestry.

**Figure S9.**
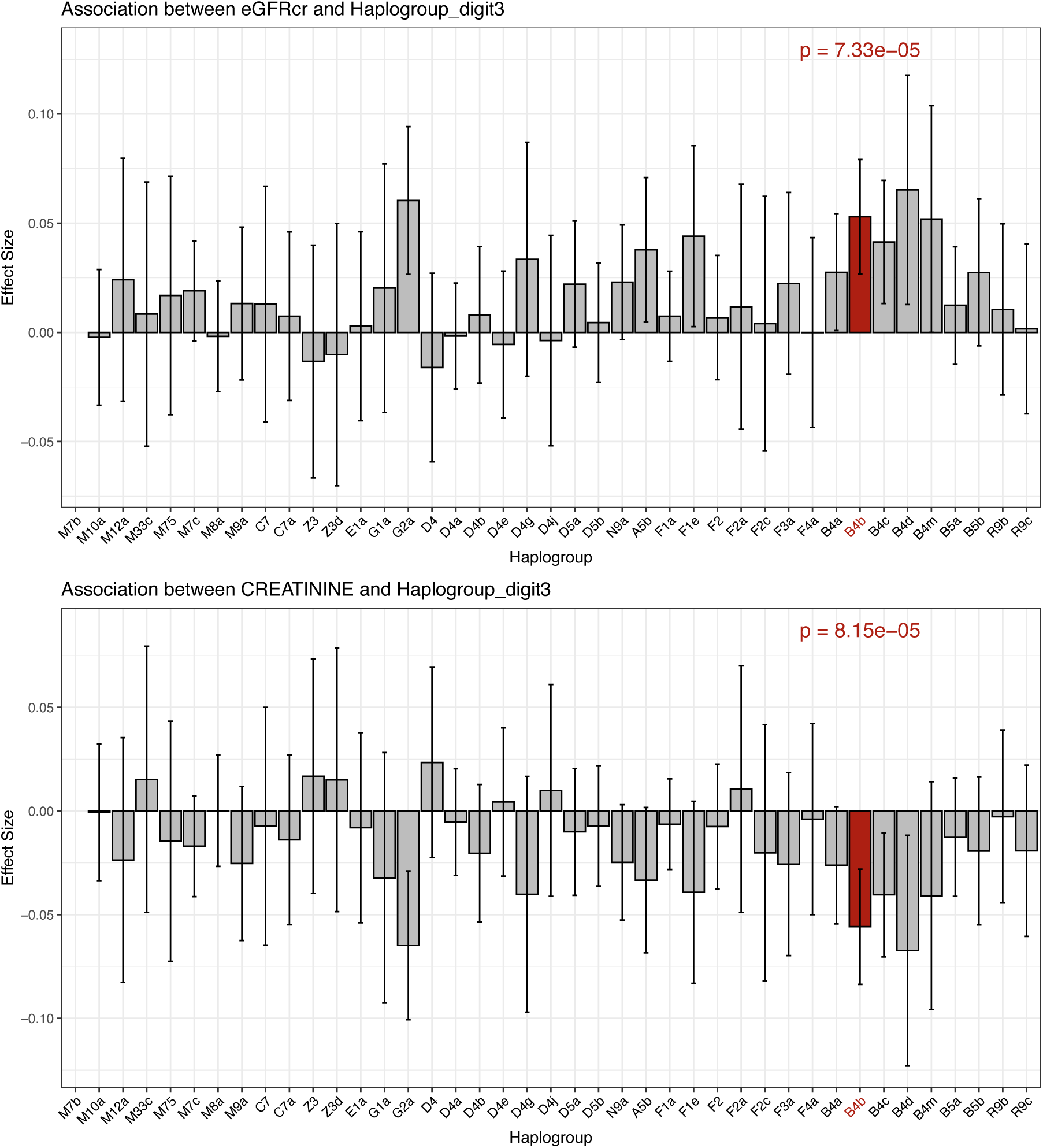
Association of mtDNA haplogroups with renal function markers. The figure displays the associations between mitochondrial haplogroups and two renal function markers: eGFR and creatinine levels. The top panel illustrates the effect sizes of each haplogroup on eGFR, while the bottom panel displays the effect sizes on Scr.

